# School Reopening Simulations with COVID-19 Agent-based Model for the Philippine Regions

**DOI:** 10.1101/2022.05.16.22275128

**Authors:** Vena Pearl Bongolan, Nico Andrew G. Francisco, Martin Thomas D. Saliba, Jimuel Celeste

## Abstract

Schools have been closed in the Philippines since March 2020 due to the COVID-19 pandemic. In 2022, the government already allowed a pilot run of limited in-person classes in low-risk areas. With such development, the present paper aims to explore the question “Is it safe to reopen schools with the current vaccination coverage?” We used an age-stratified COVID-19 agent-based model coupled with social contact probabilities to simulate school reopening and vaccination scenarios in the 17 regions of the country. Through these simulations, we found downtick points for infections and deaths—the vaccination coverage at which we do not expect increases in infections and deaths should schools reopen. We then calculated the School Reopening Viability (SRV) of the regions and visualized these scores with a stop-go map for school reopening. Simulation results suggest that all regions except Regions 7, 9, BARMM, and 13 can already reopen schools without the fear of upticks in infections nor deaths. These regions have lower vaccination coverages relative to the rest of the country, especially against the case of Luzon which has the highest vaccination coverage. We recommend that the areas of concern ramp up their vaccination efforts before reopening schools. At the same time, behavioral factors (mask-wearing, physical distancing, hand-washing) and disease resistance factors (healthy living habits) shall be enforced once schools reopen. Finally, school reopening shall be gradual to ensure the crafting of data-driven (hospital utilization, positivity rate) policies.

## I. Introduction

In March 2020, the Philippine government ordered the closure of schools due to the threat of the COVID-19 pandemic. Classes have since shifted to modular and online set-ups. With most of the student population locked in their respective households, the quality of education through online classes continues to be greatly compromised and has negative effects on their mental well-being [1]. In fact, the Asian Development Bank [2] has quantified the possible loss to the students, projecting an annual $180-worth earning loss for every student due to school closures alone. But as the pandemic continues to ease and the vaccination program continues, the Commission on Higher Education and the Department of Health jointly allowed the reopening of higher educational institutions in low-risk areas [3].

Universities and colleges can now allow students to take classes on a face-to-face basis. This is a welcome development for many as such a move will finally give students and teachers a sense of normalcy amidst the global pandemic. The high nationwide vaccination coverage is also seen as a positive indicator. As of February 2022, the Philippines already fully vaccinated 68% of its target population [4], with the National Capital Region at 79.9% of its regional population which is above the 70% target. However, caution must still be taken as new variants (Omicron) emerge as the virus continues to evolve. This context makes the calls to craft well-planned health protocols [5, 6] and to keep a gradual reopening process of schools [7] in the Philippines both timely and important. The goal is to ensure the protection of the students as well as the teachers when the classrooms finally reopen. It is a *de facto* objective for any measures to minimize infections and deaths.

In this study, we try to answer the question: Is it safe to reopen schools with the current vaccination coverage?

We used our age-stratified COVID-19 agent-based model [8] coupled with social contact probabilities to simulate various school reopening scenarios with varying vaccination coverages for the 17 regions of the Philippines. Our agent-based model is based on our previous age-stratified model [9, 10, 11] and also utilizes the social contact matrices of Prem et al. (2017) [12]. In our case, we used COVID-19 infection, recovery, and death age distributions [13] as proxy parameters for infection, recovery, and death probabilities, a method inspired by the Age-Stratification Theory that our team put forward as early as 2020 [14].

The novelty of our study lies in its context which is unique to the Philippine school reopening and the COVID-19 vaccination situation. It is an extension of our initial simulations of school reopening scenarios [15] in Quezon City, Metro Manila, the location of the University of the Philippines, which is still debating various modes of class delivery. As of this writing, there are no known studies applying modeling to simulate school reopening in the 17 regions of the Philippines. The results of the present paper bridge this gap.

## II. Methodology

### 2.1. COVID-19 Agent-Based Model

In this study, we used our age-stratified COVID-19 agent-based model (ABM) [8, 15] coupled with social contact probabilities [12]. The ABM is based on the Age-Stratified Quarantine-Modified SEIR with Non-Linear Incidence Rates Model (ASQ-SEIR-NLIR) [14, 16, 17] which is defined by the following system of differential equations:

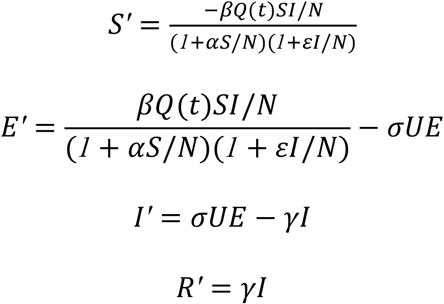

This model is a modification of the classic SEIR compartmental model. The classic model has compartments S-E-I-R representing the susceptible, exposed, infectious, and removed population. Parameters **β**, σ, and *γ*, are the transmission, incubation, and removal rates, respectively. The additional parameter Q(t) accounts for quarantine [14], U accounts for age-stratification [16], and non-linear incidence rates ⍺ and *ε* account for behavioral factors and disease-resistance factors [17]. We previously transcribed the design of this model into an agent-based model parameters and agent transition rules, resulting in an agent-based model prototype [8].

#### 2.1.1. Social Contact Probabilities

The resulting ABM was later coupled with social contact probabilities from the pre-pandemic estimates of Prem et al. [12] for the Philippines, allowing us to simulate school reopening scenarios. The social contact probabilities (Figure 1) were computed as the normalized and scaled sum of the social contact matrices (school, home, work, others). These probabilities capture the age-nuances of social interactions.

**Figure 1.**
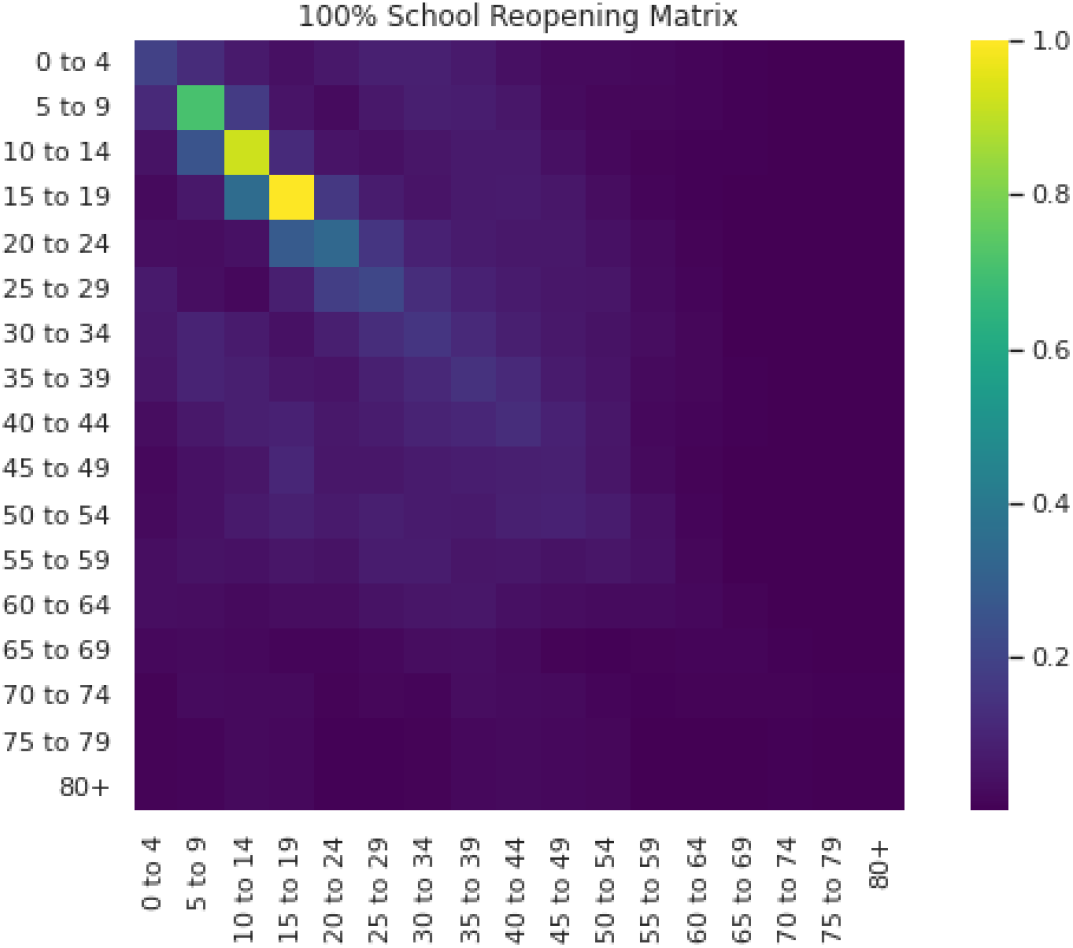
Social Contact Probabilities based on the social contact matrices of Prem et al. [12] for the Philippines. This matrix shows the 100% school reopening scenario. The diagonal signifies an apparent pattern of interaction among individuals belonging to the same age group. Interaction is highest among the younger age groups (5-24) where students usually belong.

### 2.2. School Reopening Simulations

#### 2.2.1. Vaccination and School Reopening Scenarios

In this study, we simulated the no school opening scenario (control) and the 100% school reopening scenario. The no school opening scenario reflects the situation when lockdowns are in place. On the other hand, the 100% school reopening scenario refers to the pre-pandemic situation where schools operate at 100% capacity.

We performed a vaccination sensitivity analysis for both the no school reopening and 100% school reopening scenarios. For simplicity, the control vaccination scenario was set to zero (no one is vaccinated). The other vaccination scenarios were varied by increments of 25 until 100, resulting in the following scenarios: 1) 0% (control scenario); 2) 25% vaccination; 3) 50% vaccination; 4) 75% vaccination; and 5) 100% vaccination.

In our simulations, we extracted data from March 2020 to 17 January 2022—the peak of the Omicron variant surge (36,978 new cases and 34,835 7-day average new cases). We chose this date to assume the worst case for our simulations. The datasets are cumulative, meaning the variants of concern Delta, Omicron, etc., were all captured by our simulations, along with the first or alpha-variant.

#### 2.2.2. Model Parameters

The parameters of the model are shown in Table 1. We used the parameters computed from the COVID-19 data [13] of the National Capital Region (NCR) for the infection, death, and recovery probabilities in simulating school reopening scenarios for all the 17 regions of the country instead of using the specific regional probabilities. This assumption was made as NCR has the most reported cases, owing to the concentration of testing centers and medical facilities in the region, and a theorized “street light” effect, i.e., we see the cases at the capital region where there is testing, but not in other areas where there is no testing. We take the NCR dataset as the most representative of the true distribution of infection, death, and recovery. We used these distributions as proxies for the parameters, inspired by the Age-Stratification Theory which our team put forward as early as 2020 [14].

**Table 1.** COVID-19 Agent-Based Model Parameters

| Parameters | Description | Values adopted in this study |
| --- | --- | --- |
| Infection Probabilities | Age-stratified infection probabilities | Age distribution of infections in NCR |
| Death Probabilities | Age-stratified death probabilities | Age distribution of deaths in NCR |
| Recovery Probabilities | Age-stratified recovery probabilities | Age distribution of recoveries in NCR |
| Average Incubation Period | The average number of days that an exposed individual incubates | 5 days ([18] estimated 5.6 days; we took the lower bound) |
| Average Infectious Period | The average number of days that an infectious individual infects other individuals | [19] |
| Wearing Masks | Percentage of individuals wearing masks | 100% (assumption) |
| Protection from Wearing Masks | Percentage of protection from contracting COVID-19 when wearing a mask | 65% [20] |
| Physical Distancing | Percentage of individuals observing physical distance | 60% (assumption) |
| Protection from Physical Distancing | Percentage of protection from contracting COVID-19 when observing physical distance | 90% [20] |

#### 2.2.3. SEIRDV Values

The model requires SEIRDV (susceptible, exposed, infectious, recovered, dead, vaccinated) values for every region. For simplicity, we treated each region as a single space (island-like, no ingress nor egress) and opted to use data at the regional level instead of computing the SEIRDV values for every city and/or municipality in a region (Table 2). Furthermore, each region is treated homogeneously and the sociodemographic factors were not included in the model parameters.

**Table 2.** Omicron Simulations SIRD starting values

| Regions | Susceptible |  | Infectious |  | Recovered |  | Dead |  | Vaccinated [21] |
| --- | --- | --- | --- | --- | --- | --- | --- | --- | --- |
|  | Actual | Scaled | Actual | Scaled | Actual | Scaled | Actual | Scaled |  |
| National Capital Region (NCR) | 5691316 | 49283 | 14556 | 66 | 795448 | 3104 | 10835 | 52 | 11276971 |
| Cordillera<br>Administrati<br>ve Region<br>(CAR) | 816504 | 6683 | 459 | 16 | 78464 | 314 | 1930 | 18 | 106096<br>8 |
| Region 1<br>(Ilocos) | 2411162 | 20300 | 477 | 17 | 81520 | 324 | 2042 | 18 | 323774<br>5 |
| Region 2<br>(Cagayan<br>Valley) | 1668962 | 13828 | 723 | 17 | 12641<br>8 | 500 | 4141 | 26 | 218071<br>3 |
| Region 3<br>(Central<br>Luzon) | 5635018 | 47308 | 2750 | 22 | 25729<br>5 | 1009 | 5815 | 33 | 714794<br>0 |
| Region 4-A<br>(CALABAR<br>ZON) | 7326812 | 61218 | 6050 | 35 | 45323<br>8 | 1772 | 5498 | 31 | 929971<br>3 |
| Region 4-B<br>(MIMAROP<br>A) | 1605323 | 12418 | 138 | 15 | 35624 | 147 | 1084 | 16 | 155861<br>6 |
| Region 5<br>(Bicol) | 3078416 | 1034 | 351 | 17 | 49238 | 199 | 1034 | 17 | 306029<br>6 |
| Region 6<br>(Western<br>Visayas) | 3629044 | 30487 | 535 | 17 | 11694<br>7 | 463 | 4371 | 29 | 444732<br>6 |
| Region 7<br>(Central<br>Visayas) | 3833505 | 30974 | 698 | 17 | 11779<br>3 | 467 | 5523 | 31 | 367991<br>1 |
| Region 8<br>(Eastern<br>Visayas) | 2228922 | 17511 | 275 | 17 | 45181 | 184 | 678 | 17 | 238159<br>4 |
| Region 9<br>(Zamboanga<br>Peninsula) | 1913439 | 14888 | 152 | 17 | 48070 | 197 | 1304 | 17 | 190124<br>9 |
| Region 10<br>(Northern<br>Mindanao) | 2446697 | 19257 | 269 | 15 | 73237 | 293 | 1013 | 16 | 268108<br>4 |
| Region 11<br>(Davao) | 2477678 | 20063 | 419 | 17 | 86271 | 343 | 2689 | 21 | 285124<br>0 |
| Region 12<br>(SOCCSKS<br>ARGEN) | 2397497 | 18858 | 491 | 16 | 53341 | 218 | 1100 | 16 | 216432<br>6 |
| Region 13<br>(CARAGA) | 1999044 | 16963 | 346 | 17 | 43485 | 179 | 1421 | 17 | 137222<br>5 |
| Bangsamoro<br>Autonomous<br>Region in<br>Muslim<br>Mindanao<br>(BARMM) | 2432740 | 17069 | 59 | 14 | 16812 | 72 | 491 | 17 | 854963 |

In our simulations, we assumed that there were no exposed individuals for all regions, i.e., the exposed compartment is zero. This assumption was made since there were no available data sources that would give good estimates for the number of exposed individuals. Our main data source is the Department of Health Philippines (DOH) which provided us with datasets [4, 13] where we extracted the number of infectious and removed (dead, recovered, and vaccinated) individuals.

For the removed compartment, we included all dead and recovered individuals (absolute immunity from recovery was assumed, a common simplifying assumption in epidemiological modeling). However, for vaccinated individuals, we only included a fraction: the estimated immunized. We only considered fully vaccinated individuals (full course of Pfizer, Sinovac, etc. and one shot for J&J), booster shots were not included. To estimate the number of immunized individuals, those who acquired immunity from vaccination, we multiplied the mathematical expectation for vaccine efficacy (77.08%) by the number of fully vaccinated individuals. The expected value was computed based on the vaccine supply as of February 2022 [21] and the reported efficacy of the vaccines [22, 23, 24]. This gave us the estimated number of COVID-19 immune individuals through vaccination.

Finally, for the number of susceptibles, we computed the difference between the total population (N) and the sum of exposed, infectious, recovered, dead, and vaccinated, i.e., N - (E + I + R + D + V). The regional populations were estimated from the 2020 Philippines Census [25] and the age distribution of the 2015 Census [26]. Since the age-stratified population data were not available in the latest Census, the distribution was estimated by applying the age distribution from the 2015 Census to the total regional population in 2020.

The computed SEIRDV values were then rescaled (Table 2) using an agent-to-population ratio of 1:257 to account for computational limitations. This means that highly-populated regions such as Region 4-A (CALABARZON) and National Capital Region (NCR) got more agents in contrast with lowly-populated regions such as the Cordillera Administrative Region (CAR) and Region 13 (CARAGA).

#### 2.2.4. Model Set-up

With the resulting SEIRDV and parameter values, we ran the model 20 times, with 300 model steps each run, for all the scenarios we enumerated. The results were then averaged to eliminate noise from the data.

### 2.3. School Reopening Viability

The simulation results allowed us to identify downtick points. These are vaccination coverages at which there will be no increase in infections (with respect to the control scenario: no reopening with zero vaccination) even when schools reopen 100%. We identified the downtick points for deaths separately (they are not assumed to scale!) These points were characterized as the target vaccination coverages for the regions to observe “safe” school reopening since an increase in infections, or, separately, deaths will not be expected.

With the downtick points, we then calculated the School Reopening Viability (SRV) which is the difference between the actual vaccination and the downtick points. A positive SRV means that a region can already reopen schools without the risk of infection or death increasing with respect to the control scenario. However, it should be noted that a zero SRV does not imply that there will be no more infections or deaths as schools reopen. It only implies that the situation will not become worse. A stop-go map for school reopening was then created to visualize the SRV of the regions.

## III. Results

### 3.1. Downtick Points and Vaccination Coverage

We simulated school reopening and vaccination scenarios and then computed the downtick points for infection and death. The downtick points refer to the vaccination coverage at which the change in infection or death with respect to the control scenario (no reopening with zero vaccination) is zero.

Table 3 shows the infection and death downtick points as well as the vaccination coverage of the regions as of February 2022. This allows us to identify areas with high and low downtick points as well as the areas of concern.

**Table 3.** Computed Infection and Death Downtick of Each Region together with Vaccine Coverage as of 13 February 2022.

| Regions | Infection<br>Downtick | Death<br>Downtick | Vaccine<br>Coverage<br>[4] |
| --- | --- | --- | --- |
| National Capital Region (NCR) | 17.4% | 0% | 79.9% |
| Cordillera Administrative Region (CAR) | 23.61% | 0%<sup>a</sup> | 57.8% |
| Region 1 (Ilocos) | 8.33% | 45% | 60.5% |
| Region 2 (Cagayan Valley) | 14.58% | 0%<sup>a</sup> | 58.5% |
| Region 3 (Central Luzon) | 43.41% | 42.86% | 58.4% |
| Region 4-A (CALABARZON) | 26.22% | 0% | 55.8% |
| Region 4-B (MIMAROPA) | 20.59% | 0%<sup>a</sup> | 48% |
| Region 5 (Bicol) | 45% | 0%<sup>a</sup> | 49% |
| Region 6 (Western Visayas) | 12.14% | 0% | 55.4% |
| Region 7 (Central Visayas) | 58% | 0% | 45.2% |
| Region 8 (Eastern Visayas) | 16.07% | 0%<sup>a</sup> | 49% |
| Region 9 (Zamboanga Peninsula) | 53.57% | 0%<sup>a</sup> | 49.6% |
| Region 10 (Northern Mindanao) | 55.83% | 0%<sup>a</sup> | 52.3% |
| Region 11 (Davao) | 50.81% | 0% | 52.3% |
| Region 12 (SOCCSKSARGEN) | 0% | 0%<sup>a</sup> | 42.9% |
| Region 13 (CARAGA) | 29.41% | 56.25%<sup>a</sup> | 48.8% |
| Bangsamoro Autonomous Region in Muslim Mindanao (BARMM) | 31.67% | 0% | 19.6% |

<sup>a</sup> The negative change is first achieved at this vaccine coverage but a positive change appears at a higher vaccine coverage.

First, the regions with the highest infection downtick points are Regions 7, 10, and 9. Our simulations suggest that at these points, we will not observe upticks in infections even if we reopen the schools. With 58% as the highest downtick point, we can say that the suggested targets are realistic, lower than the target vaccination coverage of the Department of Health Philippines (DOH) which is 70% for all communities.

Next, the case for the death downticks is seemingly peculiar at zero for all regions except Regions 1, 3, and 13. The regions of exception (1, 3, 13) only require up to 56.25% vaccination coverage, only slightly lower than the downtick requirement for infection, and also lower than the DOH-prescribed coverage, since infections and deaths do not scale. Meanwhile, a zero downtick point suggests that even without vaccination, deaths will not increase in these regions even if schools reopen. Particular emphasis is given to the word ‘increase’ which qualifies the concept of a downtick. The results do not suggest zero deaths but that deaths will not be worse than when schools reopen with zero vaccination. A zero downtick point shall not be treated as a default *go* sign for school reopening.

### 3.2. School Reopening Viability

With the downtick points and the vaccination coverage as of February 2022, we calculated the School Reopening Viability (SRVs) of the regions and visualized these values as a stop-go map (Figure 2). The SRVs were computed as the difference between the current vaccination coverage and the downtick points. A negative value is colored red, while a positive value is colored green, all varying according to the magnitude of the value. Intuitively, a positive SRV value means that the downtick point has already been reached; otherwise, it would be negative.

**Figure 2.**
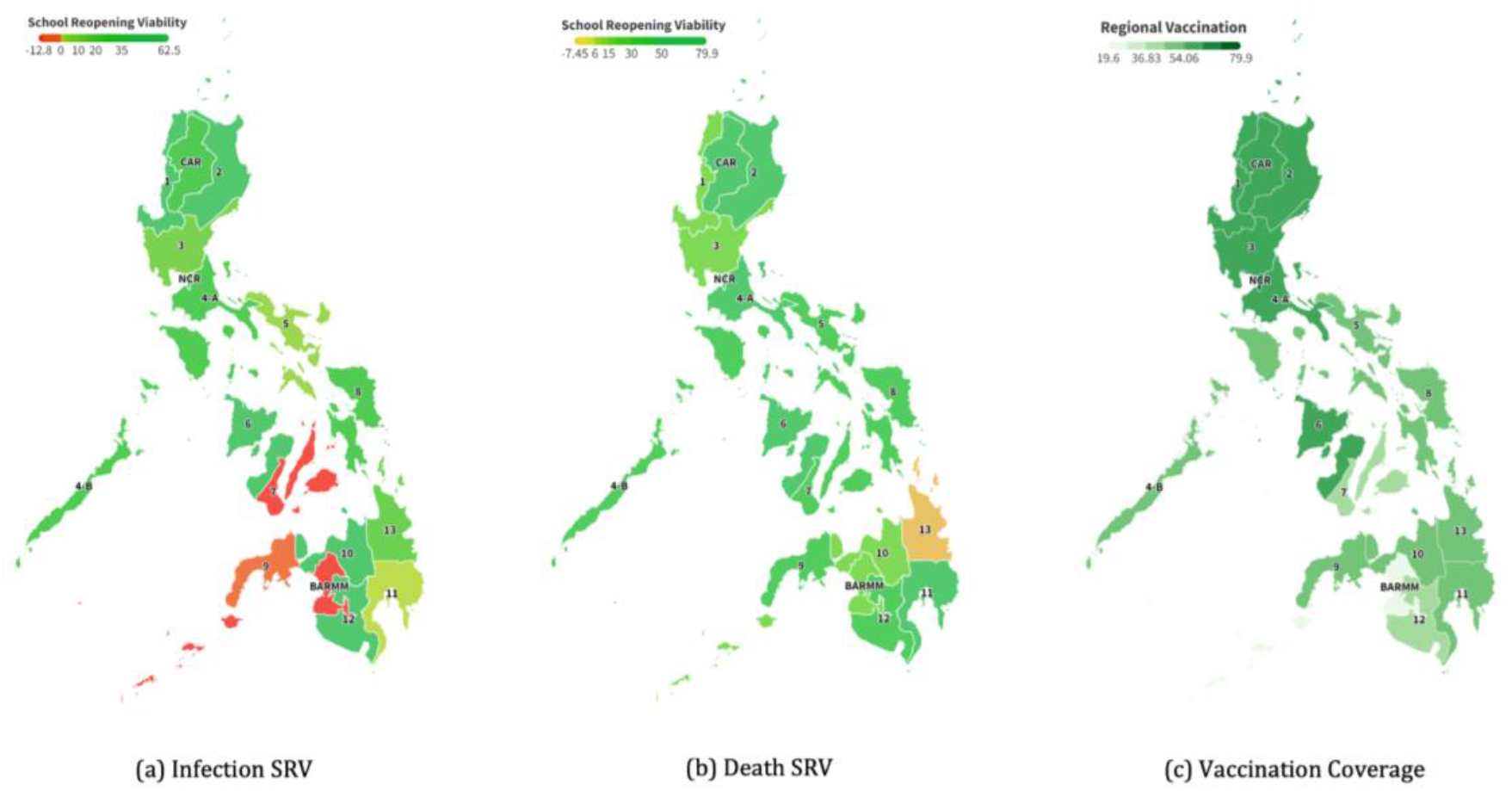
Stop-Go Map (green for go, red for stop) based on the School Reopening Viability (SRV). The SRV is an indicator that the region has already reached the downtick points (infection, death) based on their vaccination coverage. (a) Infection SRV; (b) Death SRV; (c) Vaccination Coverage as of February 2022.

As apparent from the map (Figure 2), there are four areas of concern in the southern part of the country around Visayas and Mindanao. For the infection downtick (Figure 2.a.), there are three regions of concern. These are Regions 7 (Central Visayas), 9 (Zamboanga Peninsula), and BARMM, with SRV ranging from −3.97% to −12.80%. For the death downtick (Figure 2.b.), there is only one area of concern: Region 13 (CARAGA) with an SRV of −7.45%. These values are interpreted as the additional vaccination coverage that the regions need to prevent the increase in infections and deaths when schools reopen.

The general impression with the areas of concern is that their vaccination coverage is lower than that of other places, especially compared with Metro Manila and the rest of Northern and Central Luzon (Figure 2.c.). The areas with darker greens (high vaccination) are concentrated in the northern part of the map, while the areas with lighter green (lower vaccination) are concentrated in the southern part. The disparity is apparent.

## IV. Discussion

### 4.1. Principal Results

The simulation results give us insights into which regions are areas of concern with regard to school reopening. Four such areas were identified: Regions 7 (Central Visayas), 9 (Zamboanga Peninsula), BARMM, and 13 (CARAGA). The first three regions are projected to observe upticks in infections when schools reopen, while CARAGA is projected to observe upticks in deaths. The general recommendation for these regions is to ramp up their vaccination programs to reach the downtick points (Table 3)—the points at which the infections and deaths will not increase even if they reopen the schools.

However, it shall be noted that the vaccines are never perfect as their efficacies are never perfect [22, 23, 24]. We estimated the average efficacy of the available vaccines in the Philippines to be 77.08% as of February 2022. Even if the entire population is vaccinated, we estimate that only ∼77 out of every 100 individuals would be immune from COVID-19. Reaching 100% coverage would also be impossible and is simply impractical. This is where our simulations become relevant as it provides insights regarding the “sweet spot” of vaccination for the regions. We recommend hitting these coverages first before reopening the schools but we do not recommend that they stop there until they hit the DOH-set target of 70%.

In addition to vaccination, school administrators shall also be cognizant of the importance of compliance with non-pharmacological measures such as mask-wearing, physical distancing, and handwashing. One model assumption is that individuals, including students, observe 100% mask-wearing and at least 60% physical distancing. The mask-wearing scenario is the ideal scenario while the physical distancing compliance rate is random. As we have shown in our previous study [17], these non-pharmacological measures (behavioral factors) in conjunction with disease-resistance factors (healthy living, natural immunity) are as effective as 50% quarantine in controlling the spread of COVID-19 when maximized. Hence, in line with calls for “deliberate and well-planned school health protocols [5]”, we reiterate the importance of maximizing both behavioral and disease resistance factors, especially in school settings. Our projections are only as good as the compliance of individuals to these measures since our model assumes the best behavior from the students and teachers once the schools reopen.

But this is not to say that our model is entirely optimistic. We are using a dataset [13] that covers all variants of concern (Alpha to Omicron) as of January 17, 2022, the peak of the Omicron surge in the Philippines, which is also the highest in terms of infection count. This dataset allows us to capture the worst case in terms of case incidence. Moreover, we used pre-pandemic social contact estimates [12] for school, home, work, and other locations, another way of capturing the worst-case scenario.

But then again, despite these design choices, we still think that cushioning our projections is a safer choice. We err on the side of caution. As with previous recommendations [7], the school reopening process shall be gradual. Monitoring the actual number of COVID-19 cases, hospitalization, positivity rate, and other vital safety indicators are just as important as doing simulations like what we presented in this paper. It is not enough to just meet the recommended vaccination coverage and have well-thought school protocols in place; the school reopening policies shall be data-driven to ensure timeliness, relevance, and equity.

### 4.1. Limitations

As with other modeling studies [9, 10, 11], we are limited by the quality of the data that we can access. We used the COVID-19 Data Drop of the Department of Health Philippines [13] as our main source of COVID-19 data, and so our projections are only as good as the quality of their datasets. Moreover, we devised a way to find the age distribution of the number of fully vaccinated by distributing the total according to the age distribution of the Philippine Population in 2015 [26]. Errors may have been introduced in this process but their extent is unknown.

Design-wise, our model did not capture waning immunity and so the role of vaccination booster shots was not considered. We also assumed absolute immunity from recovery (a common assumption) which can be easily incorporated as soon as we get reliable reinfection rates. Lastly, deaths from other causes (other diseases, accidents, natural) and births are not yet captured by the model. These vital statistics shall be considered in future studies.

### 4.2. Comparison with Prior Work

In contrast with other models, our age-stratified COVID-19 agent-based model coupled with social contact probabilities is closest to Covasim [11]. Ours is also age-stratified, using the social contact matrices of Prem et al. [12], and a compartmentalized model. The novelty of our study lies in its context: the Philippine school reopening and the COVID-19 vaccination situation. We utilized our agent-based model to simulate school reopening and vaccination scenarios in the 17 Philippine regions. In contrast with previous papers concerning school reopening in the Philippines [5, 6, 7], we used modeling to explore various scenarios, which to our knowledge is the first attempt.

## V. Conclusions and Recommendations

In this study, we asked: Is it safe to already reopen schools with the current vaccination coverage? We utilized our age-stratified COVID-19 agent-based model coupled with social contact probabilities to simulate school reopening and vaccination scenarios in the 17 regions of the Philippines. Our results suggest that, with the current vaccination coverage of each region and the most recent Omicron variant transmission, all regions except Regions 7, 9, BARMM, and 13 may already reopen their schools. Should school reopening be pushed in Regions 7, 9, BARMM, and 13, we believe that their respective healthcare facilities could handle the increase in infections as the regions have a 31.1%, 12.7%, and 9.3% hospital occupancy rate respectively (all still under the safe level) as of July 20, 2022 [27]. With these results, we recommend the following: 1) ramp up the vaccination in the areas of concern, 2) enforce behavioral factors in schools (mask-wearing, physical distancing, handwashing), 3) reiterate the importance of disease-resistance factors (healthy living habits), and finally 4) keep the school reopening gradually to ensure data-driven (hospital utilization, positivity rate, etc) policies moving forward. Policymakers may take insights from the study.

## Data Availability

All data used in our simulations are available online.
Our mathematical model has been well-described in previous publications. Our agent-based model is proprietary.

https://doh.gov.ph/covid-19/case-tracker

https://www.rappler.com/newsbreak/data-documents/tracker-covid-19-vaccines-distribution-philippines

## Acknowledgments

Nico thanks his family for support and Anna for her help in proofreading and providing valuable suggestions. Thomas thanks Edwin, Jess, Micko, Jeanne, Nikki, Carlo, and Vince for their support. This work is a continuation of the research on Modeling the Dynamics of COVID-19 by the System Modeling and Simulation Laboratory, and as such, we thank the past proponents of research leading to ours. We also thank DOST ASTI and COARE for the usage of their computing resources which greatly helped minimize our work. The authors acknowledge our consultants in the bigger Covid-19 team: Drs. Roselle Leah K. Rivera, Romulo de Castro, Jesus Emmanuel Sevilleja, and Salvador E. Caoili. We dedicate this work to the very worthy memory of Dr. Rogelio Pacayra Peñera of PSHS’ 81, epidemiologist, who was forcibly taken from us.

## Nomenclature

COVID-19: Coronavirus Disease of 2019
DOH: Department of Health Philippines
PSA: Philippine Statistics Authority
SRV: School Reopening Viability
WHO: World Health Organization

